# How can we make self-sampling packs for sexually transmitted infections and blood borne viruses more inclusive? A qualitative study with people with mild learning disabilities and low health literacy

**DOI:** 10.1101/2020.11.06.20217612

**Authors:** Alan Middleton, Maria Pothoulaki, Melvina Woode Owusu, Paul Flowers, Fiona Mapp, Gabriele Vojt, Rebecca Laidlaw, Claudia S Estcourt

## Abstract

**Objectives:** 1.5 million people in the UK have mild to moderate learning disabilities. Sexually transmitted infections (STIs) and blood borne viruses (BBVs) are over-represented in people experiencing broader health inequalities, which include those with mild learning disabilities. Self-managed care, including self-sampling for STIs/BBVs, is increasingly commonplace, requiring agency and health literacy. To inform the development of a partner notification trial, we explored barriers and facilitators to correct use of an STI/BBV self-sampling pack amongst people with mild learning disabilities.

**Methods:** Using purposive and convenience sampling we conducted four interviews and five gender-specific focus groups with 25 people (13 female, 12 male), with mild learning disabilities (July-August 2018) in Scotland. We balanced deductive and inductive thematic analyses of audio-transcripts to explore issues associated with barriers and facilitators to correct use of the pack.

**Results:** All participants found at least one element of the pack challenging or impossible but welcomed the opportunity to undertake sexual health screening without attending a clinic and welcomed the inclusion of condoms. Reported barriers to correct use included perceived overly complex STI/BBV information and instructions, feeling overwhelmed, and the manual dexterity required for blood sampling. Many women struggled interpreting anatomical diagrams depicting vulvo-vaginal self-swabbing. Facilitators included pre-existing STI/BBV knowledge, familiarity with self-management, good social support, and knowing that the service afforded privacy.

**Conclusion:** In the first study to explore the usability of self-sampling packs for STI/BBV in people with learning disabilities, participants found it challenging to use the pack. Limiting information to the minimum required to inform decision-making, “easy read” formats, simple language, large font sizes and simpler diagrams could improve acceptability. However, some people will remain unable to engage with self-sampling at all. To avoid widening health inequalities, face-to-face options should continue to be provided for those unable or unwilling to engage with self-managed care.

**Key messages:**

- People with mild learning disabilities found the existing self-sampling pack overly complex; many would not use it and did not feel able to engage with self-managed care at all.
- Minimum “need to know” information, very simple diagrams, and “easy read” formats specific to the needs of people with mild learning disabilities, could improve acceptability.
- Adoption of self-sampling and other elements of self-managed care without provision of alternative care models could widen health inequalities.
- Face-to-face options need to be provided but identifying those with limited health literacy will be challenging.

## Introduction

Healthcare systems worldwide are facing increasing demand.[1,2] Recent shifts in healthcare policy and practice are focusing increasingly on self-managed and remote care as a way to address demand and in some cases, increase access to care by using digital/online care pathways.[3-5] The recent COVID-19 pandemic has accelerated the pace of change.[6] Sexual health care is at the forefront of self-managed care, and services, such as postal self-sampling for STIs/BBV are commonplace[7,8] and sometimes the recommended or sole option for asymptomatic people.[7]

Self-managed care demands agency and health literacy (the ability to seek health information, understand its relevance and enable people to act on that information and make decisions).[9,10] Despite increasing use of self-managed care, the views adAcknowledgementsn opinions of people with low health literacy are not known. This is important because some of those at greatest risk of STIs/BBV are from vulnerable groups who have low health literacy and already experience considerable health inequalities, particularly regarding access and uptake of healthcare.[11]

People with learning disabilities often have low health literacy. This is because a learning disability is a lifelong condition that starts before adulthood and affects development; individuals will need assistance to understand information, learn new things and function independently.[12-14] Within the UK there are 1.5 million people with mild to moderate learning disabilities;[15,16] 26,000 of whom live in Scotland and are known to require support.[17] However, there are almost three times as many people with learning disabilities, or who have been recorded as having additional support needs when at school, who do not identify as having learning disabilities in adulthood and importantly do not benefit from learning disability services.[13]

As part of intervention development, ahead of the LUSTRUM partner notification randomised controlled trial (RCT) of Accelerated Partner Therapy (APT),[7] we explored the perspectives of people with mild learning disabilities. We aimed to identify potentially modifiable elements of the partner STI/BBV self-sampling packs with the broader aim of increasing access to this type of self-managed care for people with mild learning disabilities and people with low health literacy more broadly.

We aimed to address three research questions

1. What challenges do people with mild learning disabilities encounter with self-managed care for STIs and BBV?
2. What are the views and opinions of people with mild learning disabilities on the self-sampling pack and its contents?
3. Which elements of the pack could be adjusted to improve uptake and facilitate correct usage?

## Methods

### Participants

Participants were heterosexual people and men who have sex with men (MSM), with mild learning disabilities, aged 18-65 years and able to communicate in English.

We used existing relationships with community groups and organisations in Central Scotland and personal contacts to inform people with mild learning disabilities about the study. We sent formal letters/emails to our contacts and attended group meetings to discuss the study and answer any questions before recruitment.

### Sampling and Recruitment

We used purposive and convenience sampling, making efforts to recruit heterosexual people, MSM and people of non-cis genders.

Participant information sheets and consent forms were adjusted, to address accessibility needs (Appendix 1 & 2). The lead researcher, AM, is a registered learning disability nurse. His expertise was used to develop all recruitment materials and other elements of the research processes which needed adapting.

We recruited participants through gatekeepers (charitable/voluntary organisations) or directly, depending on the preference of the community group. Where recruitment was via gatekeepers we provided an information sheet which detailed inclusion criteria and the participant information sheet to enable selective distribution. Inclusion criteria and ability to give informed consent were initially assessed by gatekeepers. Where we sought consent directly from potential participants, we assessed comprehension and understanding of the study information, and ability to use this to make an informed decision. We also assessed ability to give informed consent before and throughout the interview/focus group.

Interviews and focus groups took place at community settings in Central Scotland between July and August 2018. Participants were required to dedicate approximately one hour to the study and had the option of a support person being present for part or all of the research activities. Participants were compensated with a £30 voucher.

All interviews were audio recorded using digital devices and were transcribed in a Word document format for the purpose of analysis. Data collected were fully anonymised for reporting, presentation, archiving and/or publication purposes. All data were securely transported, transferred and stored in compliance with relevant data management guidelines.

In total, we conducted four interviews with one male and three female participants, and five focus groups comprised of three all-male groups with a total of 11 participants and two all-female groups with a total of 10 participants (Table 1).

**Table 1.** Participant Characteristics

|  | **Gender** | **Female** | **Male** |
| --- | --- | --- | --- |
| **Age** | 18-25 | 1 |  |
|  | 26-35 | 5 | 5 |
|  | 36-45 | 4 | 5 |
|  | 46-55 | 2 | 2 |
|  | 56+ | 1 |  |
| **Ethnicity** | White Scottish | 12 | 12 |
|  | Asian/Asian British | 1 |  |
| **Relationship Status** | Single | 3 | 7 |
|  | Cohabiting (living with another person) | 5 | 2 |
|  | Married | 1 | 2 |
|  | Separated/Divorced | 1 |  |
|  | Widowed | 1 |  |
|  | Other (in a relationship but not cohabiting) | 3 | 1 |
| **Employment Status** | Part-time employed | 2 | 4 |
|  | Unemployed | 9 | 8 |
|  | Retired | 1 |  |
|  | College course | 1 |  |
| **Sexuality** | Heterosexual | 9 | 9 |
|  | Bisexual |  | 1 |
|  | Prefer not to say | 4 | 2 |

### Procedure

The researchers, AM, GV and RL, conducted the interviews and focus groups utilising a semi-structured topic guide (Appendix 3). Following introductions, the researcher(s) provided an explanation of the study. Researchers reminded participants that the interview/focus group would be audio-recorded and they were free to leave at any point and explained processes for data handling and ensuring anonymity and confidentiality.

The researchers checked participants’ understanding of the purpose of the interview or focus group, answered any questions and then sought informed written consent and completed demographic questions.

We began the interview/focus group by showing the participant(s) either the male and female self-sampling/APT packs and discussing how it should be used (Figure 1). We explained that the packs would be provided to people diagnosed in clinic with chlamydia (described as the most common STI) to take to their sex partners. For this to happen, the sex partner would first need to have a telephone consultation with a qualified healthcare professional in clinic. We also explained that we had already conducted extensive pre-trial work with diverse groups of people recruited from sexual health clinics and community settings to develop the packs’ contents and instructions.

**Figure 1:**
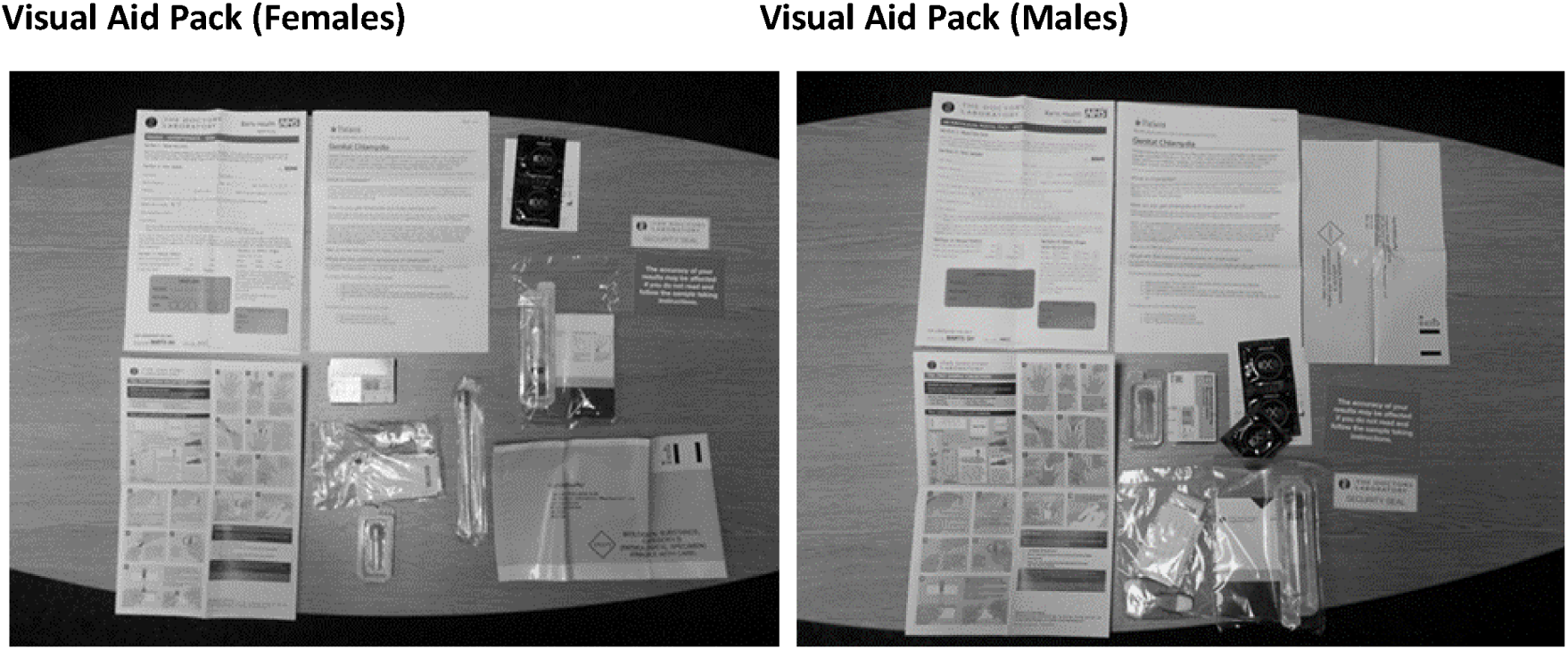
Self-sampling packs used in interviews and focus groups. Legend: The packs contained information about chlamydia; instruction leaflet on how to use the pack; urine collection container (for males) or vulvo-vaginal swab (for females); blood sample collection kit; return envelope; security seal; red label (advising accuracy of tests depends on following sample taking instructions) and completing the laboratory form, and condoms.

The topic guides were used to elicit barriers and facilitators to using the pack before exploring the testing kit contents and how they are used.

We asked participants to open the packs, look at the contents and tell us what they thought of the packs. We then asked participants to read the instructions in the pack, describe the contents of the pack and explain how they would use the contents.

### Analysis

We combined focus group and individual interview data. We balanced deductive and inductive thematic analyses of audio-transcripts[18] to explore issues associated with the barriers and facilitators to correct use of the pack and its contents. Deductive analysis focussed a priori on the barriers and facilitators to the correct use of the pack and its contents. In this way, the analysis balanced an inductive understanding of how participants understood the contents of the self-sampling pack and its use. Firstly, we conducted an initial sweep of the data enabling us to code in relation to the main elements of the pack, corresponding to the key research questions. Then we conducted a second round of more participant-led, inductive analysis. AM then coded these broad elements of the transcripts to generate participant-led themes. AM & MP reviewed and discussed the themes before defining and naming them. The data generated were stored and organised using NVivo software (Version 11).

Ethical approval was obtained from Glasgow Caledonian University School of Health and Life Sciences Ethics Committee (HLS/PSWAHS/17/194).

## Results

Six key themes emerged. Table 2 shows representative verbatim quotations illustrating the underlying themes, relating to our three research questions.

**Table 2.** Extracts illustrating themes within participants’ accounts

| **Research question 1: What challenges do people with mild learning disabilities encounter with self-managed care for STIs and BBV?** |  |
| --- | --- |
| **<u>Emergent Themes</u>** | **<u>Illustrative Quotations</u>** |
| **Accessing sexual health care** | *"If you've got learning difficulties, you need the help. You can't just read that..." [leaflet]. GF1*<br>*"Well, you've explained it [self-sampling pack] to me so it's easy when somebody's explaining to me." DF1*<br>*"...but if you don't have a comprehension about why you'd be getting this [self-sampling pack], so that would freak you out." GF3* |
| **Support from others** | *"...if you live at home, with no support, and you don't want your mum to know that you're sexually active, how do you go about it?" GF3*<br>*"I wouldn't ask somebody that I couldn't trust because I would like to keep that private." DF1*<br>*"I'd get my support worker to help me." EM2*<br>*"I'd rather go to the doctor's, 'cause then you'd know what's getting done, right then." GF4* |
| **Research Question 2: What are the views and opinions of people with mild learning disabilities on the self-sampling pack and its contents?** |  |
| **Using the pack** | *"...it's not giving you, like, instructions, like it's not a clear indication there of how to use it." DF2*<br>*"Well, to be honest with you, it can be a bit daunting." DM4*<br>*"Are these for likes of to find out if you've got sexual diseases as well...as well as doing it the other way? Because I've not heard of doing it* |
|  | *this way.” DF1* |
| **Accessibility of the pack** | *“...the steps, the diagram is okay, but the writing should be a [little] bit bigger.” DM4*<br>*“...because it [stages on leaflet] goes across, and down, is that confusing, would it be easier if it had everything in a row?” GF3* |
| **Research Question 3: Which elements of the pack could be adjusted to improve uptake and facilitate correct usage in this group?** |  |
| **Contents of the pack** | *“Yeah, but it's not explaining it more, see if it's done the right way or the wrong way... so it's not clear... I think that would be a lot tricky for some people to get caught out on.” GM5*<br>*“But what, why, what STI is it for? [self-sampling pack]” GF8*<br>*“But it's like, they've gave you a blood sample bottle, but they've given you nothing to take it with.” GM3*<br>*“Definitely include them [condoms] because people might not want to get infected again.” DF3* |
| **Using the contents of the pack** | *“I'm not going to say what I think... I just call it my back passage...See, you wouldn't know if that's the back to the front... [anatomical diagram]” DF1*<br>*“Because I think you could do the swab, and you might have taken it wrongly. Or you could have taken it incorrectly, and it would have given an improper reading.” GF3*<br>*“These [blood sample kit] look like what, if you're a diabetic, you have to go and get your sugars done, and that's what I was meaning.” GF1*<br>*“I'm diabetic, so I know I'm used to needles.” GM3* |

**Legend:** Due to the small sample size, participant characteristics are limited to gender to prevent deductive disclosure (F: female, M: male, G/D/E code relates to locality)

We present a narrative discussion of each theme, drawing out barriers and facilitators to using the pack and its contents.

### Accessing sexual health care

This theme identified some of the significant challenges that participants experience, particularly when accessing and trying to understand new and complex information.

Participants’ knowledge of STIs was limited and this compounded the challenges of grasping new information. Although the participants all had a mild learning disability, this encompassed a range of cognitive abilities, specific difficulties and literacy skills. Written information was thought to be particularly challenging or inaccessible by all.

### Support from others

Many participants explained that decisions about their health and wellbeing are often undertaken by others and restrictions put on risk taking behaviours. Some participants continue to live with parents, highlighting this as a particular difficulty for sexual health when privacy was important.

Most participants received some supports in their daily lives and often relied on guidance from others when navigating uncertain and unfamiliar areas. This was often with someone they trust and where privacy is respected. Which was also the case when faced by a self-sampling pack.

Participants voiced a need for someone else to help navigate the pack and due to the sensitivity or privacy around sexual health issues this was an additional consideration when asking for help. For some, the complexity of the pack and the knowledge and understanding required to undertake self-sampling meant that they would rather go to their general practitioner (GP) or sexual health service than try themselves.

### Using the pack

Most participants described feeling overwhelmed to varying degrees when opening the pack and did not know where to start. This could prevent them proceeding further.

Most participants found the detail included in the chlamydia information sheets (the infection, health consequences, treatment and partner notification) (Appendix 4) to be too long and difficult to read. They could not relate the information to the actual sampling kits in the pack.

Despite the challenges voiced by most of the participants, the opportunity to use a self-sampling pack at home was welcomed by some due to convenience. Some also perceived self-sampling less embarrassing than attending a sexual health clinic or GP.

### Accessibility of the pack

The inclusion of diagrams and pictures were seen as a welcome step towards an easier to read format by all participants. However, participants voiced problems interpreting the diagrams which illustrated the anatomical sites for self-sampling. This was a particular problem for women who had difficulties relating the diagrams to their own anatomy. Written information relating to each of the tests contained in the pack was felt to enhance the usability of the pack.

Participants suggested several improvements to aid clarity and remove ambiguity. These included adopting an “easy read” format,[19] avoiding columns of text, and simplifying how the key health messages are presented within the pack to create a more user-friendly feel. Specific suggestions included making it easier to identify items mentioned in the guidance notes with the pack components by numbering them and cross-referencing. For some, an accompanying online “YouTube” video would be welcomed. (Videos were available as part of the wider trials however participants taking part in this study were not made aware of them).[20]

### Contents of the pack

The number of test components in the pack created some anxiety and participants had difficulty understanding their purpose. The perceived lack of a clear process and sequence for undertaking the different activities needed to successfully self-sample was problematic. Interpreting anatomical diagrams depicting sampling sites and diagrams showing what to do with the samples were thought to be particularly challenging.

Condoms contained in the pack were familiar to most participants and were seen as a positive step in preventing future STIs.

### Using the contents of the pack

Overall participants found the process daunting and at times, confusing. They voiced fears about efficacy, most stating that they would need support to undertake the tests. Obtaining samples was felt to be particularly difficult; most participants felt unclear about what was required, how to take the samples and what to do with them subsequently.

Many women did not seem to have sufficient understanding of their own anatomy, and experienced difficulties in interpreting the anatomical diagrams. This lead to a lack of confidence in their ability to follow the instructions provided to take a vulvo-vaginal swab. They also voiced concerns about appropriate technique and the potential for issues with reliability of the test by doing it incorrectly.

The motor skills and manual dexterity required for taking blood samples gave cause for concern and were felt to be a significant barrier to successful self-sampling. However, where participants had previous experience of similar procedures, such as diabetic monitoring, the familiarity gave more confidence.

## Discussion

All participants in this study found at least one element of the pack challenging or impossible. Some felt able to manage some of the tasks alone or with help from someone trusted but others expressed bewilderment and a lack of ability to engage with the pack. However, many welcomed the opportunity to undertake sexual health screening without attending a sexual health clinic and the inclusion of condoms in the pack was regarded as a positive STI prevention message.

Blood sampling and interpretation of female self-swabbing instructions appeared particularly difficult. Reduced manual dexterity, low knowledge of genital anatomy and overly complex instructions and diagrams contributed to the problems described. Adjustments to the pack structure and contents might increase accessibility, including limiting text to only the minimum required for completing self-samples, “easy read” formats, simple language, large font sizes and simpler diagrams of anatomy and how to take samples.

To our knowledge, this is the first study to explore barriers and facilitators to use of an STI/BBV self-sampling pack in people with learning disabilities. As sexual ill health disproportionately affects those with limited agency, low literacy and low health literacy, these findings increase our understanding of the challenges of self-managed care in this vulnerable group and suggest how problems might be overcome.

There are several limitations. Our sample was restricted to people from one area of Scotland and is unlikely to represent the full range of learning disabilities people experience. Most self-managed sexual health care requires initial digital engagement, such as ordering a pack online, whereas our study focussed “downstream” on the pack itself. In reality, it is likely that some people would not have managed to take even the first step of ordering online and therefore effectively excluded from self-managed care pathways altogether. We had already extensively optimised the packs by simplifying their components in the context of pre-trial development work for a partner notification randomised controlled trial.[7] Self-sampling packs in contemporary use may not have been developed with this focus on usability. As such, our findings may somewhat overestimate acceptability and feasibility of this type of care for people with mild learning disabilities.

Evidence suggests that uptake of online postal self-sampling is greater in people from more affluent areas,[21] which may be viewed as a proxy for health literacy level. It is perhaps unsurprising that people with mild learning disabilities perceived considerable barriers. Evaluations of existing online self-sampling services report sample return rates of 54 to 72.5 %; and blood samples are the least likely to be returned.[22,23] Blood sampling was felt to be particularly challenging by our participants, partly due to difficulties with manual dexterity often coexisting with learning disabilities. Participants with prior experience of diabetic monitoring perceived fewer barriers which suggests that opportunities to learn how to collect blood samples might encourage engagement.

A previous qualitative study of young people’s perceptions of smartphone-enabled self-testing and online care for STIs, suggested that some participants were concerned about the accuracy of self-testing.[21] Similar views were expressed in this study but concerns related to the participants’ perceived ability to take appropriate samples rather than the accuracy of the test itself.

Although the development of the packs was informed by extensive qualitative research with the intention of creating a simple-to-use pack, and to fit the financial constraints of the trial, it was not accessible to most of our participants. This suggests that people with even mild learning disabilities and people with low literacy, will need highly tailored self-sampling packs to enable engagement with this type of self-managed care.

“Easy read” formats,[19] specific layouts, reduction and simplification of information to the minimum, avoiding ambiguity and having a step by step guide, with a clear start and end point for sequencing of tests, and ensuring kit components are clearly matched to the accompanying instructions could facilitate pack use. An accompanying video clip could be helpful but this would need to accurately reflect the exact pack the person is using to avoid further confusion.

Even if adjusted packs are provided, there will be considerable challenges in identifying who would benefit from them, as many people with mild learning disabilities and people with poor health literacy are not known to sexual health services. People from these groups may be more likely to take inadequate samples or sample incorrectly which may lead to false negative results due to poor technique rather than true absence of infection.

As sexual health care becomes increasingly self-managed, we risk excluding vulnerable individuals because they are unable or unwilling to engage with processes which require sophisticated levels of health and digital literacy.[13] Education and training for health care professionals to assist identification of those individuals is likely to help but an “online first” approach[24] may exclude people with mild learning disabilities from engaging with care at all. Easily accessible alternatives (face-to-face options) are essential to avoid widening health inequalities further.

Although we focussed on self-sampling packs for STIs & BBV our findings will have broad generalisability to other areas of healthcare. These findings may also be of use with people from a broad range of ethnicities, people who speak English as a second language or who do not speak English.

Sexual health services have been at the forefront of self-managed care but little attention has been given to the needs of those who are not health and digitally literate. Future work needs to quantify and characterise people with sexual health needs and risks who choose not to, or are unable to engage with self-managed care. To avoid amplifying health inequalities further we must develop ways of identifying people who may need additional support in a timely and sensitive way and develop accessible alternative models of care.

## Additional Files

**Appendix 1:** Participant information sheet

**Appendix 2:** Consent form

**Appendix 3:** Semi-structured interview/focus group schedule

**Appendix 4:** Chlamydia information sheets

## Data Availability

The data that support the findings of this study are available from the corresponding author, [AM], upon reasonable request.

## Acknowledgements

We would like to thank all of the participants who took part in focus groups and interviews and all individuals and organisations who helped with recruitment for this study. This study has been shaped through ongoing discussion and support from the LUSTRUM team (Claudia S Estcourt (Principal Investigator), Alex Comer, Alison R Howarth, Andrew Copas, Anna Tostevin, Anne M Johnson, Catherine H Mercer, Chidubem (Duby) Ogwulu, Christian Althaus, Fiona Mapp, Gabriele Vojt, Jackie Cassell, Jean McQueen, John Saunders, Karen Pickering, Maria Pothoulaki, Melvina Woode Owusu, Merle Symonds, Nicola Low, Oliver Stirrup, Paul Flowers, Rak Nandwani, Sarah Lasoye, Sonali Wayal, Susannah Brice, Tracy Roberts) and the Programme Steering Committee (PSC) (Simon Barton (chair), Alex Miners, Artemis Koukounari, David Crundwell, Emmanuel Rollings-Kamara, Lynis Lewis, Rebecca Turner, Robbie Currie, Rachel Shaw, Saima Saddiqui (on behalf of NIHR programme management).

