## Appendix 1 for "How can we make self-sampling packs for sexually transmitted infections and blood borne viruses more inclusive? A qualitative study with people with mild learning disabilities and low health literacy"

### Appendix 1: “Testing a sexual health treatment pack”

#### Participant Information Sheet

**Study 5**

#### Group discussions with participants from the public

We would like to invite you to take part in a research study on improving a sexual health service. We would like to hear what you think about a sexual health treatment pack. Before you decide it is important you understand what taking part in the study will involve. Please read the following information carefully and discuss it with others if you wish.

**Who we are**

We are researchers working at Glasgow Caledonian University and University College London. We are currently working on a study called LUSTRUM, which aims to improve sexual health services in the UK. The study is funded by the National Institute for Health Research (NIHR).

**Why are we doing this study?**

Sexual health is very important, and is part of the wider public health. In recent years, there has been an increase in infections which can be passed on during sex (sexually transmitted infections or STIs). It is very important that people who have a sexually transmitted infection (STI) get treated, so that they get better and the infection does not spread to their sex partners. This is why we would like to find out what people think about a new sexual health treatment pack. This treatment pack is for the sex partners of people who have an STI.

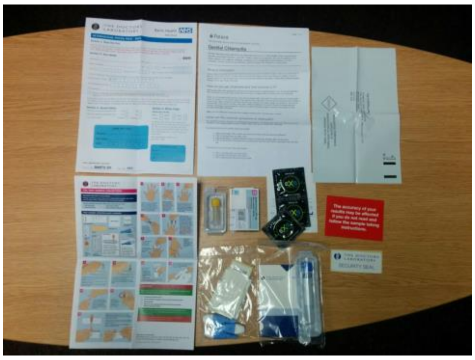

This is what the sexual health treatment pack contains. We will show you the treatment pack on the day of the group discussion.

**Why have we invited you to take part?**

We want many different people to be involved in our study. We invite people who are 18-65 years old and who are able to decide for themselves if they want to take part in the study.

**Do you have to take part?**

You do not have to take part in the study if you don’t want to. If you decide to take part, we will call you to arrange the day and time to talk with us whenever it is best for you.



GCU Glasgow Caledonian University
Funded by NHS National Institute for Health Research

**What will happen if you decide to take part in the study?**

If you agree to take part, we will ask you to take part in a group discussion with other people and the researchers. We will show you the treatment pack and then we will ask you questions to find out what you think about it, and if you think it is easy or difficult to use and why you think that. We will not ask you anything personal.

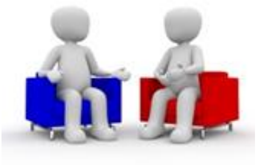

There is no right or wrong answer to these questions. We just want to know what you think. We will also give you a questionnaire with some questions about your age, ethnic background, employment, education level, sexuality, and relationships.

Before we start the group discussion we will ask you to sign a consent form, which means you agree to take part in the study. You can bring a member of staff, a relative or a friend to the group discussion if you wish.

#### How long will the group discussion last?

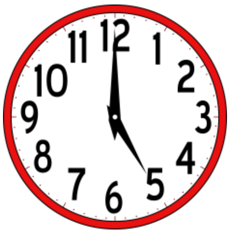

The group discussion will last approximately 1 hour, and this is a one-off discussion, so we will not ask you to come back.

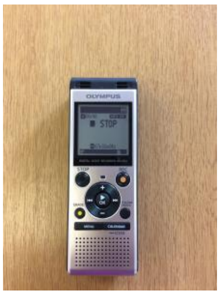

We will record the group discussion to help us remember everything you say.

#### How will we keep the recording safe and make sure you are not identified from it?

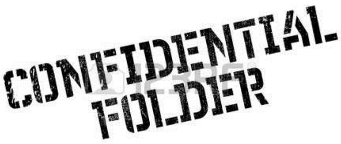


Your personal details will be kept private. We will not share with other people personal things you say to us, unless it has to do with keeping you safe. In this case, we will tell someone who can help you like your support worker, or your doctor.

#### What are the possible advantages of taking part?

Taking part in this study will not help you personally. As a way to thank you for your time and sharing your thoughts with us, we will give you a £30 voucher after the group discussion. You can use the voucher in most high street shops (e.g. Boots, Argos, Iceland etc).

#### What are the possible disadvantages of taking part?

We do not expect any disadvantages but during the group discussion we will check if you feel comfortable and happy to answer questions. You can decide to leave the group discussion at any time without giving us a reason.

#### What will happen to the results of the study?

The results of the study will be shared with healthcare services, academics, professionals and practitioners, community based organisations and service users. We will use the things you and others say to write reports and academic papers, and make presentations about the study. We may use the exact words that you say but we will not use your name. Instead we will use numbers or made up names

#### What if you want more information?

We will share updates of the research and its progress through the project website (www.lustrum.org.uk) and through the project twitter account ‘@LUSTRUM_5’

If you want more information about the study you can contact:

**In Glasgow:**

Dr Maria Pothoulaki
Glasgow Caledonian University


Professor Paul Flowers
Glasgow Caledonian University


Mr Alan Middleton
Glasgow Caledonian University


**Thank you for taking the time to consider taking part in this study**
