## Appendix 2 for "How can we make self-sampling packs for sexually transmitted infections and blood borne viruses more inclusive? A qualitative study with people with mild learning disabilities and low health literacy"

### Appendix 11: "Testing a sexual health treatment pack"

#### Study 5

#### Group discussion - Consent Form

Researchers: Dr Maria Pothoulaki, Mr Alan Middleton, Professor Paul Flowers (Glasgow Caledonian University)

**Please tick the boxes below if you agree**

I have read and understood the information sheet about the study (dated 1 September 2017, version 1). ☐

I have had the opportunity to ask questions ☐

My age is between 18 and 65 years of age ☐

I understand that if I want I can leave the group discussion at any point ☐

GCU Glasgow Caledonian University

Funded by NHS National Institute for Health Research

I agree to the group discussion being audio-recorded ☐

I agree for researchers to use my direct words (quotations) in publications, reports and/or presentations, without revealing my name. ☐

I understand that my personal details will not be shared outside the research team ☐

I understand that the only reason for sharing my details is if I am in danger and need help to keep me safe ☐

I understand the information about the study and I can decide to take part ☐

I agree to take part in the study ☐

| ______________ | ______________ | ______________ |
|---|---|---|
| Please write your name | Date | Please sign above here |

| ______________ | ______________ | ______________ |
|---|---|---|
| Researcher's name | Date | Researcher's signature |
