## Appendix 3 for "How can we make self-sampling packs for sexually transmitted infections and blood borne viruses more inclusive? A qualitative study with people with mild learning disabilities and low health literacy"

### Appendix 3: "Maximising intervention reach to wider populations: Testing the APT intervention pack"

#### INTERVIEW SCHEDULE

**Study 5**

#### I. Introduction

- *Introduction of the facilitator/researcher and participant*
- *Explain information about the study, the purpose of the interview and the type of questions that will be asked*
- *Explain that the participant is free to leave at any point*
- *Explain the use of the audio-recorder and how data will be handled*
- *Explain what type of information/data will be shared and with whom (anonymity and confidentiality)*
- *Check understanding of information, ask if participant has any questions and take informed written consent*
- *Ask Demographic questions*

  *(Audio-recorder on)*

#### II. Presentation of the Accelerated Partner Therapy (APT) intervention pack

- *Presentation of visual aid (APT Pack) and a brief background summary of how it will be used*

#### III. Assessing the strengths and weaknesses of the APT pack

- *Ask participant to open the pack, have a look at the contents of the pack and voice their thoughts*
- *Ask participants to read carefully the instructions in the pack*
- How do you think about the pack?
- What are the contents in this pack?
- How are the contents used?



Funded by NHS National Institute for Health Research

*Example of questions to be asked for each of the items in the APT pack*

- Why is the Genital chlamydia information placed in the pack?
- What is good about it?
- What is bad about it?
- How could it be better?

*(Ask the above questions for each of the APT pack items: condoms, instruction leaflet on how to use the pack, medication, urine collection container, blood collection pack, envelope, security seal, red label, laboratory form.)*

- If you were to use the pack, what would you find most difficult?
- If you were to use the pack, what would you find most easy?
- What/who would help in using the pack?
- What would you recommend to the people who make the pack?

#### IV. Conclusion

*Check if the participant has any questions or comments.*

Thank you for participating in our study!
