## Appendix 4 for "How can we make self-sampling packs for sexually transmitted infections and blood borne viruses more inclusive? A qualitative study with people with mild learning disabilities and low health literacy"

Patient

View this article online at: patient.info/sexual-health/sexually-transmitted-infections-leaflet/chlamydia

### Chlamydia

Around 3 to 7 in 100 sexually active young people in the UK are infected with chlamydia. It is most common in those aged under 25.

#### What is chlamydia?

Chlamydia is a sexually transmitted infection (STI) caused by a germ (bacterium) called *Chlamydia trachomatis*. In women, chlamydial infection usually affects the neck of the womb (cervix) and the womb (uterus). In men, it usually affects the pipe through which urine is passed (the urethra) in the penis. Chlamydia also sometimes causes infection of the eye, throat and lungs.

#### How do you get chlamydia and how common is it?

59%

of people would be worried about contracting chlamydia from a casual sexual encounter.

Source: Patient Sexual Health Survey (https://patient.info/sexual-health-at-christmas)

Most people with chlamydia get the infection by having sex with an infected person. You can become infected with chlamydia if you come into contact with the semen or vaginal fluids of a person who has chlamydia.

In 2015 in England, there were 200,288 new cases of chlamydia. Around 3 to 7 in 100 sexually active young people in the UK are infected with chlamydia. It is most common in those aged under 25. Many of those infected have no symptoms - 7 in 10 infected women and 5 in 10 infected men. They would not be aware they have the infection. You can be infected with chlamydia for months, even years, without realising it. The risk of infection increases with the number of changes of sexual partner. Wearing a condom during sex helps to protect you from chlamydia and other STIs.

**Note**: you **cannot** catch chlamydia from hugging, kissing or from sharing cups or cutlery.

#### What are the common symptoms of chlamydia?

##### Symptoms in women

No symptoms occur in around 7 in 10 infected women. Also, you may not have any symptoms until several weeks (or even months) after coming into contact with chlamydia.

If symptoms do occur in women, they may include:

- Vaginal discharge. This is due to the neck of the womb (cervix) becoming inflamed.
- Pain or burning when you pass urine.
- Vaginal bleeding or spotting between periods. In particular, bleeding after you have sex.
- Pain or discomfort in the lower tummy (abdomen) area (the pelvic area), especially when you have sex.

##### Symptoms in men

No symptoms occur in 5 in 10 infected men.

If symptoms do occur in men, they may include:

- Pain or burning when you pass urine.
- Discharge from the end of your penis.
- Pain or discomfort at the end of your penis.

### Do I need any tests?

##### Tests for females

The best test is a vaginal swab. A swab is a small ball of cotton wool on the end of a stick which is used to obtain mucus and cells to send to the laboratory for testing. It is inserted about 5 cm into your vagina and turned (rotated) gently for a few seconds. This test can be done by a health professional but it is also possible for you to do the test yourself. This is usually an option you are given if invited to have screening tests for chlamydia.

This is the most accurate test. There are two other possible tests. One is a swab taken from the neck of the womb (cervix) by a nurse or doctor. The other is a urine test. When testing urine for chlamydia you should provide a urine sample after not having passed urine for at least an hour. You catch the first part of the urine stream in the container.

##### Tests for males

For men the usual test is a urine test, collected in the same way as for women above. The other option is for a health professional to take a swab from the pipe through which urine is passed (the urethra) in the penis.

##### Other tests

If you have had anal or oral sex then you may have a back passage (rectal) or throat swab taken.

If infection with chlamydia is confirmed, you will be advised to have tests for other STIs.

**Note**: the **cervical screening (cervical smear) test** does **not** test for chlamydia.

### What is the treatment for chlamydia?

It is important that treatment for chlamydia should be started without delay. In some people where chlamydia is strongly suspected, this may even mean starting treatment before test results are available. Prompt treatment reduces the risk of complications in the future.

A short course of an antibiotic medicine usually clears chlamydial infection. You should tell your doctor if you are (or may be) pregnant or are breast-feeding. This may affect the choice of antibiotic. You should not have sex until you and your sexual partner have finished treatment (or for seven days after treatment with a single-dose antibiotic).

The most commonly used antibiotics are:

- Doxycycline one tablet twice a day for seven days; **or**
- Azithromycin one tablet only - single dose.

Other options are used if these are not suitable for you. Your doctor will advise.

### Does my partner need to be treated?

Yes. Also, any other sexual partners within the previous six months should also be tested for infection.

If your sexual partner is infected and not treated then chlamydia can be passed back to you again after you are treated.

There may be certain occasions when you may not want to contact partners from previous relationships. In these cases staff at the clinic can contact previous partners for you without disclosing your details. This is because it is important that anyone who is at risk of infection with chlamydia be both identified and treated.

### Why should I have treatment if I have no symptoms?

If you are infected with chlamydia, it is essential that you take treatment even if you do not have any symptoms of chlamydial infection. Reasons for this include:

- The infection may spread and cause serious complications (see below). This can be months or years after you are first infected.
- You can still pass on the infection to your sexual partner(s) even if you do not have symptoms.

### Do I need to be tested again after treatment?

You do not usually need to have a test to check the treatment worked if you have taken an antibiotic medicine correctly. However, it is advisable to have another test for chlamydia in the following situations:

- If you think you have had sex with a person with chlamydia.

- If your symptoms do not improve after treatment.
- If you had unprotected sex before you finished the treatment.
- If you did not complete the course of treatment.
- If you are pregnant. (If you are pregnant and have been treated for chlamydia, you should have another test three weeks later.)

Also in England, the national screening programme advises that if you are aged under 25 and have had a positive test for chlamydia, you should have a repeat test three months later. This is to check the infection has cleared completely and that you have not got it back again.

#### What are the possible complications of chlamydia?

- If left untreated, the infection may seriously affect the womb (uterus) and Fallopian tubes - this is called pelvic inflammatory disease (PID). 10-40 women in 100 with chlamydia develop PID. This may develop suddenly and cause a high temperature (fever) and pain. It can also develop slowly over months or years without causing symptoms (also known as silent PID). However, over time, scarring or damage to the Fallopian tubes may occur and can cause:
    - Persistent (chronic) pelvic pain.
    - Difficulty becoming pregnant (infertility).
    - An increased risk of ectopic pregnancy if you become pregnant. In this condition, the pregnancy develops in a Fallopian tube and can cause serious life-threatening problems.

- The risk of developing some complications of pregnancy, such as miscarriage, premature birth and stillbirth, is increased in pregnant women with untreated chlamydia.
- If you have untreated chlamydia during childbirth, your baby may develop a chlamydial infection of their eye or lung during the birth.
- Possibly reduced fertility in men.
- Reactive arthritis is a rare complication which can occur both in men and in women. In this condition, you get painful swollen joints. It may also appear as combined symptoms of inflammation of the eye and of the pipe through which urine is passed (urethra). It may be due to the immune system 'over-reacting' to chlamydial infection in some cases.

The risk of complications is much reduced if chlamydial infection is treated early.

#### Who can be screened for chlamydia?

In England there is a National Chlamydia Screening Programme. This offers chlamydial screening for sexually active women and men aged under 25 years. In this age group, screening is undertaken yearly or each time these women and men have a new sexual partner. The aims of this programme are to detect chlamydia early so it can be treated promptly. This should reduce the risk of transmission and also reduce the risk of developing complications. You can find information about screening at your GP surgery or local pharmacy. It is also available through family planning clinics, genitourinary medicine (GUM) clinics or online.

In countries where there is not a screening programme, testing is still offered regularly to sexually active young people. You can request testing regularly if you are in this category. You can do this through your GP or by attending a GUM clinic. It may be available in other ways (for example, online) depending on the area in which you live.

Certain other groups of people are also recommended to undergo screening for chlamydia. For example:

- If you have a partner with chlamydia.
- If you have another STI.
- If you are a semen or egg donor.
- If you are having an abortion (termination of pregnancy).
- If you have had two or more sexual partners in the past year.

Men will be asked to give a urine sample and women can either give a urine sample or take a swab. A swab is a small ball of cotton wool on the end of a stick, used to take a sample of mucus and cells for laboratory testing. Women can take the swab themselves from the lower vagina.

#### Further reading & references


- Sexually transmitted infections (STIs): surveillance, data, screening and management; Public Health England, 2016
- Sexually Transmitted Infections in Primary Care; Royal College of General Practitioners and British Association for Sexual Health and HIV (Apr 2013)
- National guideline for the management of chancroid; British Association of Sexual Health and HIV (2014)
- Towards elimination of HIV transmission, AIDS and HIV-related deaths in the UK; Public Health England (November 2017)


**Disclaimer:** This article is for information only and should not be used for the diagnosis or treatment of medical conditions. Patient Platform Limited has used all reasonable care in compiling the information but makes no warranty as to its accuracy. Consult a doctor or other healthcare professional for diagnosis and treatment of medical conditions. For details see our conditions.

| | | |
|---|---|---|
| Author:<br>Dr Laurence Knott | Peer Reviewer:<br>Dr John Cox | |
| Document ID:<br>4216 (v43) | Last Checked:<br>16/10/2017 | Next Review:<br>15/10/2020 |

View this article online at: patient.info/sexual-health/sexually-transmitted-infections-leaflet/chlamydia

Discuss Chlamydia and find more trusted resources at Patient.

Patient Access

Book appointments,
order repeat prescriptions and
view your medical record online

To find out more visit
www.patientaccess.com
or download the app

GET IT ON Google Play

Download on the App Store

© Patient Platform Limited - All rights reserved.
